# A transcriptomic approach to understand patient susceptibility to pneumonia after abdominal surgery

**DOI:** 10.1101/2023.01.25.23284914

**Authors:** Hew D Torrance, Ping Zhang, E Rebecca Longbottom, Yuxin Mi, Justin P Whalley, Alice Allcock, Andrew J Kwok, Eddie Cano-Gamez, Cyndi G Geoghegan, Katie L Burnham, David B Antcliffe, Emma E Davenport, Rupert M Pearse, Michael J O’Dwyer, Charles J Hinds, Julian C Knight, Anthony C Gordon

## Abstract

**Rationale:** Nosocomial infections are a major healthcare challenge, developing in over 20% of patients aged 45 or over undergoing major-abdominal surgery, with postoperative pneumonia associated with an almost five-fold increase in 30-day mortality.

**Objectives:** To describe immune-pathways and gene-networks altered following major-abdominal surgery and identify transcriptomic patterns associated with postoperative pneumonia.

**Methods and Measurements:** From a prospective consecutive cohort (n=150) undergoing major-abdominal surgery whole-blood RNA was collected preoperatively and at three time-points postoperatively (2-6, 24 and 48hrs). Twelve patients diagnosed with postoperative pneumonia and 27 matched patients remaining infection-free were identified for analysis with RNA-sequencing.

**Main Results:** Compared to preoperative sampling, 3,639 genes were upregulated and 5,043 downregulated at 2-6hrs. Pathway-analysis demonstrated innate-immune activation with neutrophil-degranulation and Toll-like-receptor signalling upregulation alongside adaptive-immune suppression. Cell-type deconvolution of preoperative RNA-sequencing revealed elevated S100A8/9-high neutrophils alongside reduced naïve CD4 T-cells in those later developing pneumonia. Preoperatively, a gene-signature characteristic of neutrophil-degranulation was associated with postoperative pneumonia acquisition (*P*=0.00092). A previously reported Sepsis Response Signature (SRSq) score, reflecting neutrophil-dysfunction and a more dysregulated host response, at 48hrs postoperatively, differed between patients subsequently developing pneumonia and those remaining infection-free (*P*=0.045). Analysis of the novel neutrophil gene-signature and SRSq scores in independent major-abdominal surgery and polytrauma cohorts indicated good predictive performance in identifying patients suffering later infection.

**Conclusions:** Major-abdominal surgery acutely upregulates innate-immune pathways while simultaneously suppressing adaptive-immune pathways. This is more prominent in patients developing postoperative pneumonia. Preoperative transcriptomic signatures characteristic of neutrophil-degranulation and postoperative SRSq scores may be useful predictors of subsequent pneumonia risk.

## Introduction

Every year in high-income countries, 1 in 10 adults undergo a non-cardiac surgical procedure (1), with a mortality rate in Europe as high as 4% (2). In those aged 45 or over, surgical resection remains the mainstay of treatment for solid organ malignancies, accounting for the bulk of major abdominal surgical procedures performed in this age range. Postoperative nosocomial infections remain a significant cause of morbidity and mortality (3). Over 20% of surgical patients aged 45 or older suffer clinically significant infectious complications (3, 4), with a patient who develops postoperative pneumonia having an almost five-fold increased risk of 30-day mortality compared to those without infection (3).

Pneumonia risk is amplified following major abdominal surgery as patients often exhibit residual effects from general anaesthesia, opioids and occasionally neuromuscular blockade (5). Several strategies, including postoperative continuous positive airway pressure have failed to reduce pneumonia incidence (5).

The temporal changes in immune function following major abdominal surgery are complex (6, 7) with our understanding largely extrapolated from the response to polytrauma (8, 9), major joint replacement (10) or from previous reductionist approaches, assaying alterations to single, or relatively small combinations of cytokines, correlated to clinical outcome (11, 12). From these data it is currently unclear why only some individuals develop postoperative infectious complications despite a similar intraoperative insult and baseline condition.

Cancer itself drives wide-ranging immunological alterations that facilitate immune tolerance, thus accelerating tumour growth and dissemination (13), with parallels drawn between this disease and what is observed in protracted infection (14, 15). The influence of damage-associated molecular patterns (DAMPs) released by tissue damage from major surgery in patients already experiencing chronic DAMP-mediated stimulation due to active malignancy, is largely unexplored. Indeed, patients with active cancer are often excluded from analysis, despite this being a highly clinically relevant, vulnerable and extensive patient cohort.

Distinct sepsis response signatures (SRS) derived from white blood cell transcriptomic profiling have been previously described (16-18), associated with differences in immune function and outcome (17, 19, 20). It is now increasingly apparent that there are parallels between the immune pathways activated following exposure to circulating pathogen-associated molecular patterns (PAMPs) and DAMPs (21) and that it may be helpful to delineate shared and distinct biological processes between infectious and sterile inflammation (22).

In this study we used a systems-biology approach to define changes in immune pathways and gene networks activated in whole blood samples collected before and after major abdominal surgery in a well-phenotyped cohort of predominantly cancer patients. We aimed to first, systematically describe transcriptomic patterns following major abdominal surgery and second, identify differential patterns of gene expression and SRS/SRSq that are associated with the risk of postoperative nosocomial pneumonia.

## Methods

The BIONIC study was a prospective observational study recruiting patients aged 45 or over undergoing elective major abdominal surgery. Ethics approval was granted by the East Midlands – Nottingham 2 Research Ethics Committee (14/EM/1223). The inclusion/exclusion criteria and recruitment process are described in detail in the **Supplementary Methods**. Data were collected on each patient until hospital discharge and included information on co-morbidities, American Society of Anesthesiology (ASA) physical status classification, indication for surgery, cancer staging and diagnosis, duration of the procedure, intensive care unit admission and in-hospital mortality. All patients received standardised perioperative prophylactic antibiotic therapy and were examined daily for the presence of infection. Definitions of infection were agreed prospectively by the investigators and were based on the Centre for Disease Control and Prevention (CDC) criteria (23).

### Blood sampling and patient selection

PAXGene (Qiagen, USA) RNA tubes were collected immediately before induction of anaesthesia (preoperatively) and then 2-6, 24 and 48hrs postoperatively. Pared EDTA samples were drawn and a differential white cell count performed.

Patients with core sample sets (preoperative, 24 and 48hrs postoperatively, n=86) were identified from the total BIONIC cohort (n=150). Of these, twelve patients subsequently developed a postoperative pneumonia. In a pragmatic manner, taking into account RNA-sequencing cost and statistical power (24), 27 patients with similar clinical characteristics to the pneumonia cohort, who remained infection free were selected as comparators. Samples obtained at 2-6hrs postoperatively were available for 11 of the 39 patients (total 128 samples for RNA-sequencing). The clinical characteristics considered for selection were based on key factors that could affect patient postoperative immune competence including surgery type, operation duration, smoking status, presence of diabetes, existing co-morbidities and laparoscopic surgery. The patient characteristics are outlined in **Table 1 & Supplementary Table E1**, and in the STROBE diagram (**Figure 1**).

**Table 1:**
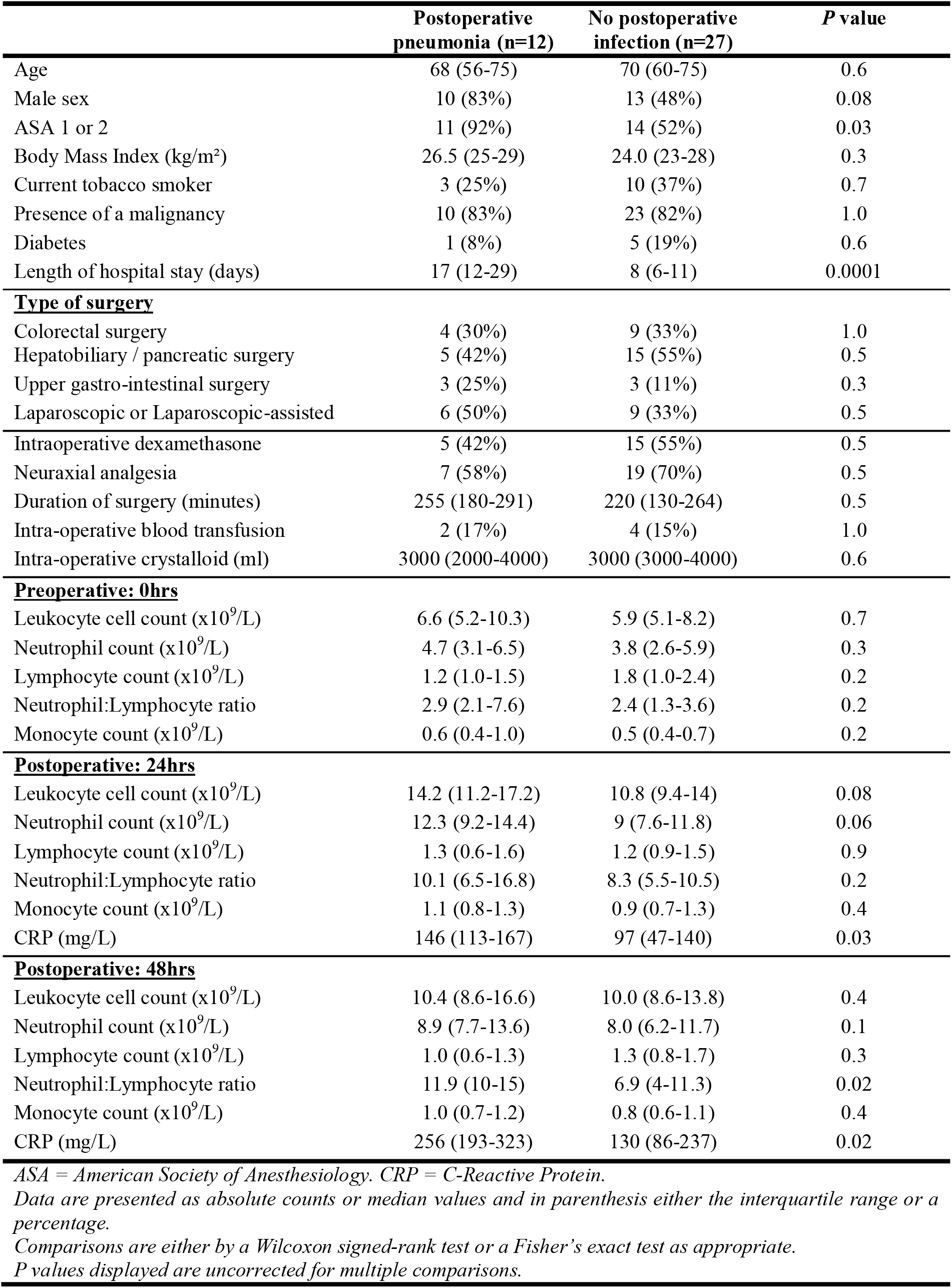
BIONIC transcriptomics cohort patient characteristics

**Figure 1.**
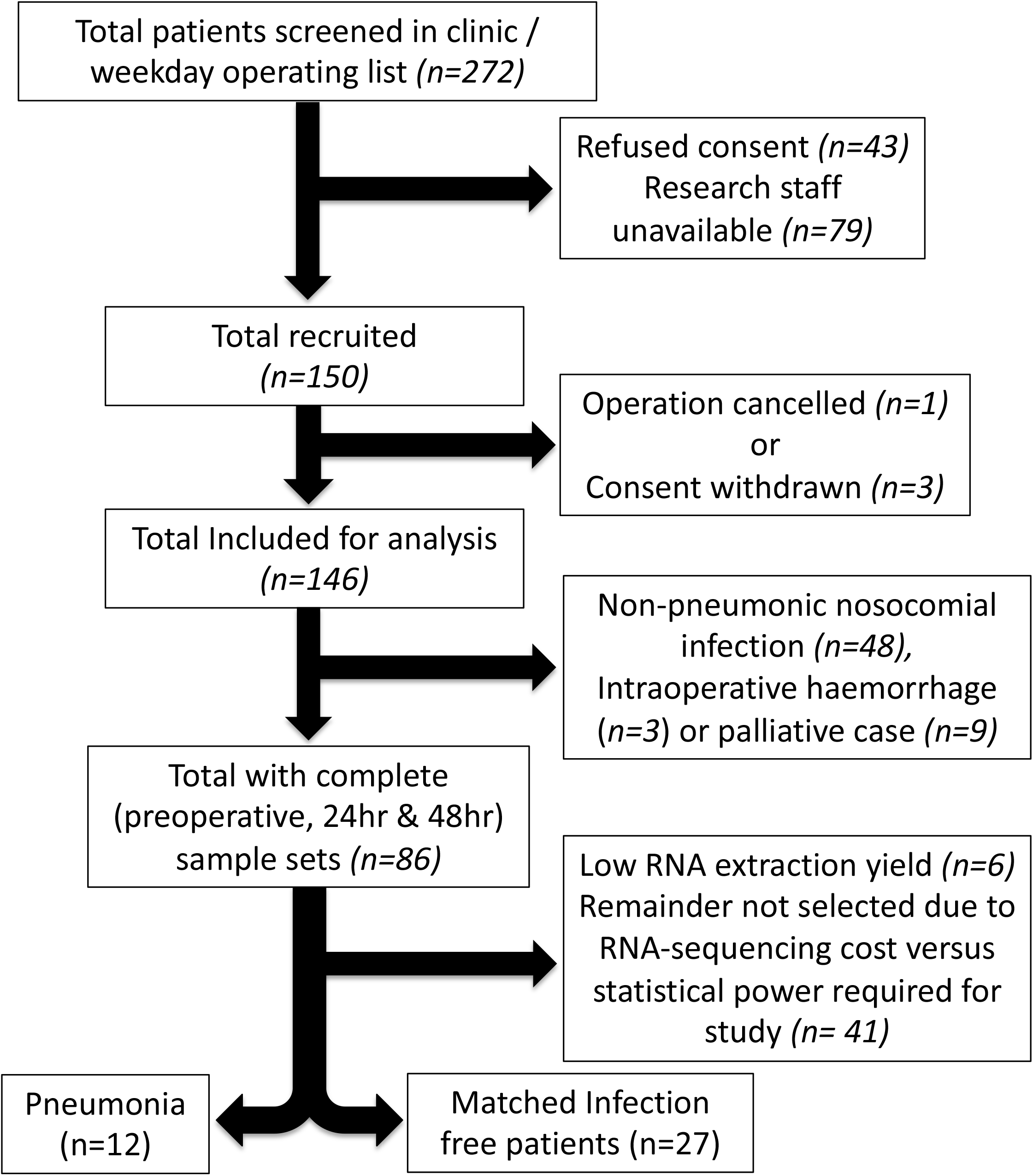
A STROBE diagram outlining the patient selection process for transcriptomic analysis. Patients (n=12), with complete sample sets, who suffered a postoperative pneumonia were identified for analysis. A cohort (n=27), who had remained infection free, were identified as comparators.

### RNA-Seq data analysis

RNA sequencing, alignment, determination of gene counts and gene differential expression was described in detail in the **Supplementary Methods**. Two samples were removed during QC. All downstream analysis was performed using R v4.0.3. R packages fgsea, and MSigDB v7.4.1 including Canonical pathways (KEGG, BIOCARTA, REACTOME, PID and WikiPathways) and Gene Ontology (GO) terms were utilised for gene-set enrichment analysis (GSEA) by hypergeometric test, while cell-type deconvolution was performed using CIBERSORTx (25). Patient clusters were defined by hierarchical agglomerative clustering based on a similarity measure (Euclidean distance) and Ward’s method or k-means (Hartigan-Wong algorithm). Performance of the gene signatures was evaluated in caret (v6.0.92). Area Under Curve (AUC) and Receiver Operating Characteristic (ROC) curves of the models were calculated and plotted using MLeval (v0.3).

## Results

### Acute transcriptomic response in patients following major abdominal surgery and signatures of subsequent postoperative pneumonia

Longitudinal RNA-seq analysis of whole blood (**Fig. 2A and Fig. E1A**) from 39 patients enabled us to capture the evolution of the transcriptomic response to surgery as well as distinct signatures between patients who did, or did not, subsequently develop a postoperative pneumonia (occurring median six [IQR 5-9.5] days postoperatively). Principal component analysis (PCA) illustrated that preoperative samples clustered together (grey dots; **Fig. 2B**) and are clearly distinct from the postoperative samples (**Fig. 2B**). There were acute transcriptional changes comparing 2-6hr following surgery to preoperative samples (red vs. grey circles), with a gradual return to baseline in 24hr (cyan) and 48hr (purple circle) samples. Furthermore, clear differences were observed between 48hr samples in those who would later be diagnosed with pneumonia vs. infection free patients (**Fig. 2B $ Fig. E1B** solid vs. dashed circle). Transcriptomic differences were not observed when comparing other clinical features including sex, age, duration of surgery and smoking status (**Fig. E1C**). At 2-6hr postoperatively, 3,639 genes were upregulated while 5,043 were downregulated, compared to preoperative samples (fold change > 1.5, FDR < 0.05; **Supplementary Table E2**). Pathway analysis demonstrated innate-immune activation including changes in neutrophil degranulation and Toll-like-receptor (TLR) signalling (**Fig. 2C**) alongside adaptive immune suppression, characterised by downregulation of CD28-mediated T-cell activation and increased expression of T-cell exhaustion markers (**Fig. 2C; Supplementary Table E3**).

**Figure 2.**
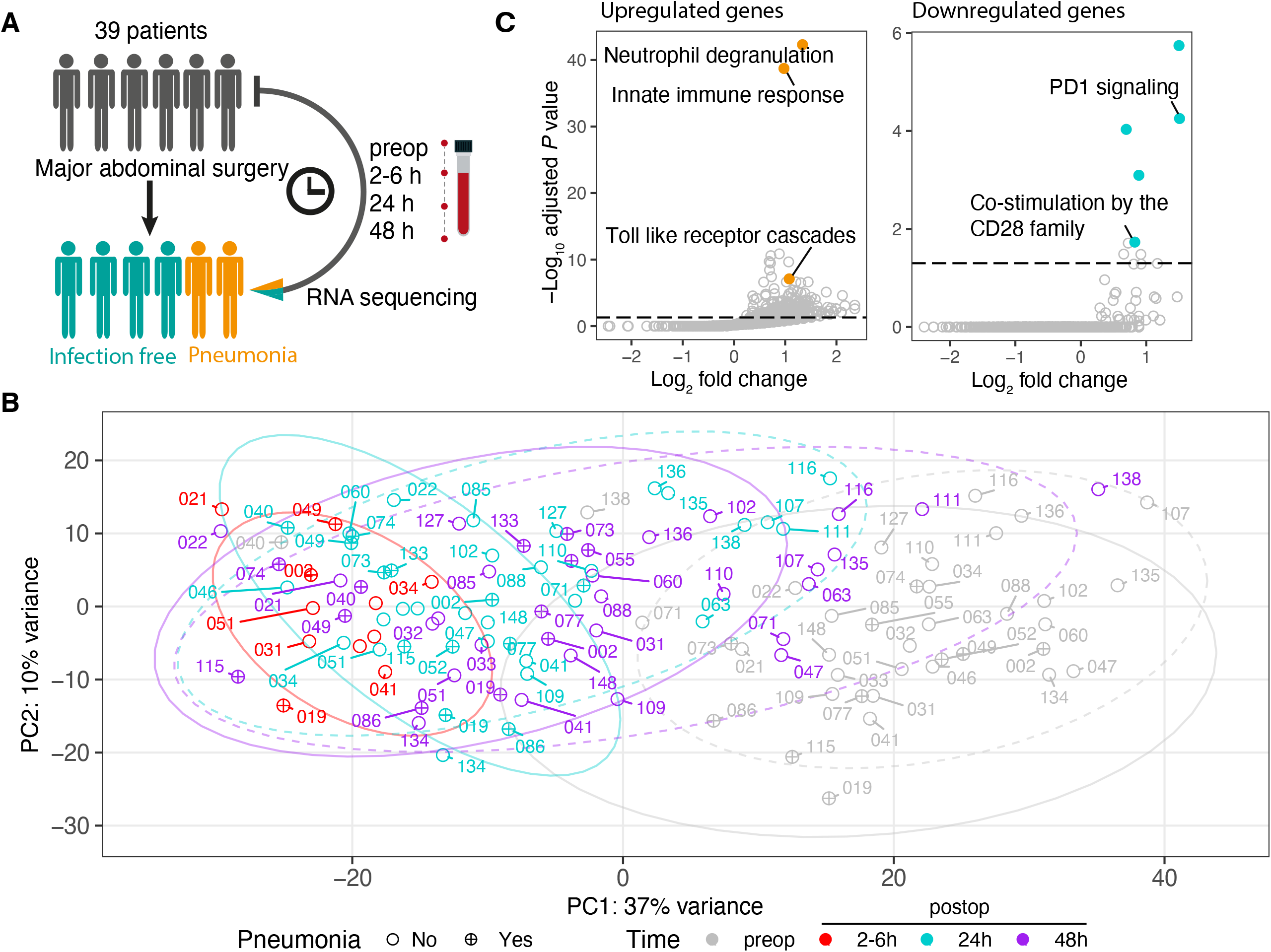
Sustained acute response in major abdominal surgery patients is associated with later postoperative pneumonia diagnosis. (A). Schematic diagram of a longitudinal RNA-seq analysis of whole blood isolated from patients undergoing major abdominal surgery. (B). Principal component analysis of gene expression in samples preoperatively and postoperatively in 2-6, 24 and 48hrs. Each dot represents a sample, labelled with patient ID. Colours indicate different time-points. Shapes indicate the clinical status of patients after operation (circle: infection-free; circle plus: pneumonia). Ellipses indicate a 95% confidence interval for the groups (dotted line: infection-free; continuous line: pneumonia). (C). Scatter plots showing the fold enrichment of upregulated differentially expressed genes or downregulated genes (2-6h vs. Preop) on the x-axis (log_2_ scale), and the adjusted *P* value on the y-axis (-log_10_ scale) in pathways annotated by Reactome. Hypergeometric enrichment analysis was performed to identify pathways in which DE genes were overrepresented (see **Supplementary Methods**). The horizontal dashed line represents the Benjamini-Hochberg-adjusted *P* value 0.05.

### Relative lymphopenia and differences in S100A8/9-high neutrophils, and hematopoietic stem and progenitor cells in patients who develop postoperative pneumonia

Next, we sought to determine temporal cell-type alterations, specifically exploring the differences between patients ultimately diagnosed with postoperative pneumonia compared to those remaining infection-free. When considering the hospital laboratory leukocyte differential counts generated concomitantly with transcriptomic sampling (see **Supplementary Methods**), innate immune cells including neutrophils and monocytes increased postoperatively, alongside a reduction in lymphocytes (**Fig. E2A**). No significant differences in hospital laboratory cell count were seen between patients developing pneumonia and those remaining infection-free (**Fig. E2B**), although the neutrophil:lymphocyte ratio (NLR) was increased at 48hrs in those who developed pneumonia (*P*=0.02), (**Table 1**). To detect if differences in the relative frequencies of cell populations occurred at a more granular level, cell-type deconvolution using CIBERSORTx (25) was performed on the bulk RNAseq samples (**Fig. 3A**; see **Supplementary Methods**). Preoperative samples from patients who ultimately developed a postoperative pneumonia demonstrated reduced numbers of naïve CD4 T cells and B cells, and significantly elevated numbers of mature neutrophils, hematopoietic stem and progenitor cells (HSPCs) and S100A8/9-high neutrophils (**Fig. 3B; Supplementary Table E4**). These differences in naïve CD4 T cells, HSPCs and S100A8/9-high neutrophils were also present postoperatively at 24 and 48hrs (**Fig. 3C; Supplementary Table E4**).

**Figure 3.**
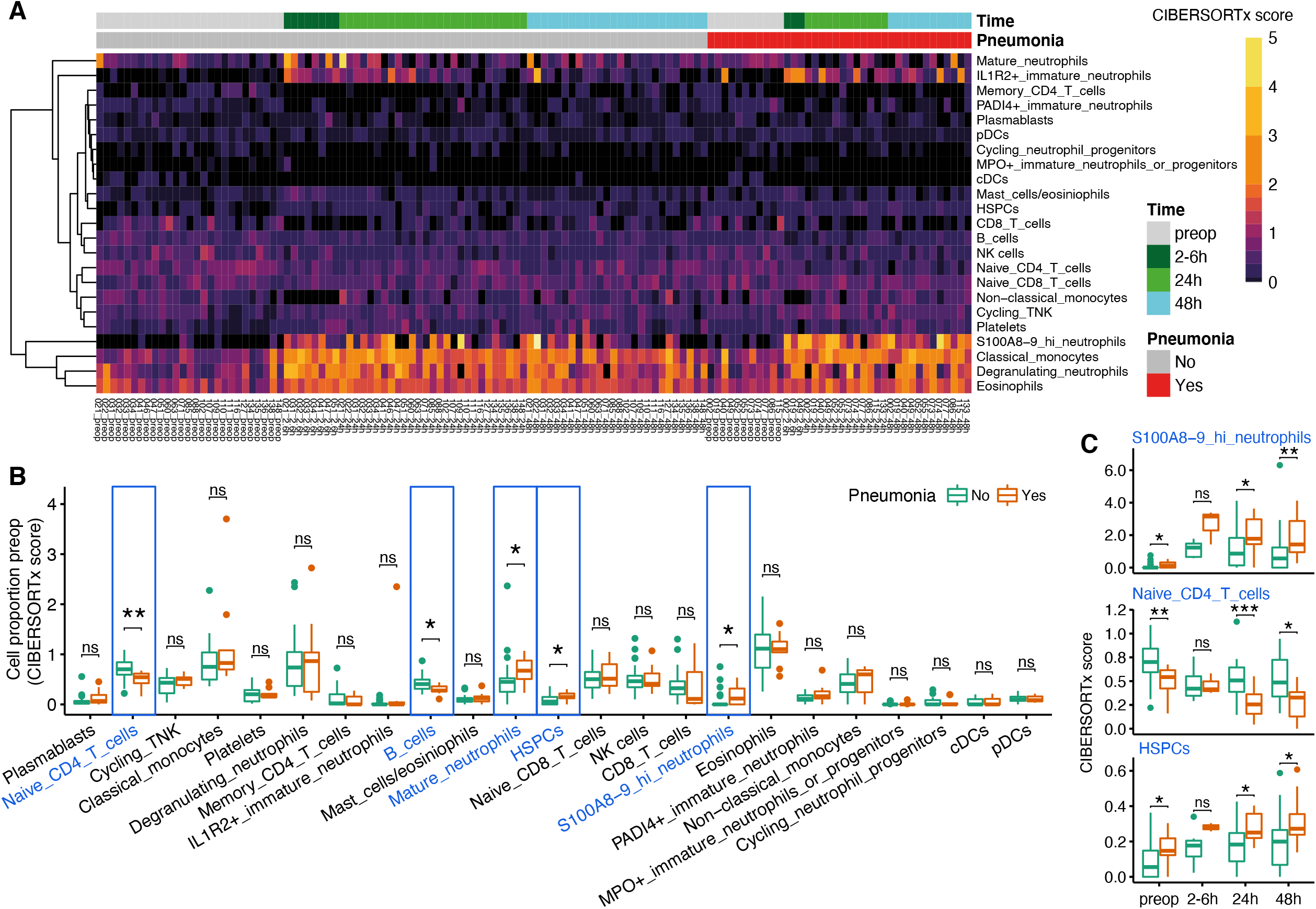
Specific cell populations and pathways showing differential abundance and enrichment before and after abdominal surgery, and relationship with subsequent development of pneumonia. (A). Heatmap of CIBERSORTx absolute scores of 23 different cell types in 126 bulk RNA-seq samples. The absolute proportion of each cell type (CIBERSORTx absolute scores) was obtained from deconvolution using a single cell RNA-seq reference panel (see **Supplementary Methods**). (B). Box plots outline the proportions of 23 different cell types in preoperative samples from patients who did (orange) or did not develop pneumonia (cyan). *P* value was calculated by two-tailed Wilcoxon signed-rank test. * *P* < 0.05, ** *P* < 0.01, *** *P* < 0.001, *n.s*. = not significant. (C) Box plots of CIBERSORTx absolute scores of indicated cell types in samples from patients who did (orange) or did not develop pneumonia (cyan) across different time-points after surgery.

Because the incidence of cancer in this cohort was more than 80%, we assessed the influence of this diagnosis on our findings. No significant association was detected between the presence of cancer and a diagnosis of postoperative pneumonia (**Fig. E2C**). No differences were identified in the abundance of naïve CD4 T cells, HSPCs or S100A8/9-high neutrophils at any sample time-point in those patients with or without a cancer diagnosis (**Fig. E2D**).

### Differential gene expression and pathway enrichment in patients developing postoperative pneumonia observed before and after surgery

We subsequently sought to identify differences in specific genes and pathways, both pre and postoperatively that were associated with postoperative pneumonia. The number of genes differentially expressed between pre-operative and post-operative samples over time reduced in patients remaining infection-free (**Fig. 4A**; bar plot upper panel). In contrast, the number of differentially expressed genes was sustained in patients who later developed pneumonia (**Fig. 4A**; bar plot middle panel). This difference is reflected in the principal component analysis (**Fig. 2B)**. The number of genes differentially expressed between those patients that would develop pneumonia compared to those that would remain infection-free was greater preoperatively (131 genes) and at 48hrs (581 genes) than during the more acute phase (2-6 or 24hrs) (**Fig. 4A** & **4B; Supplementary Table E5**).

**Figure 4.**
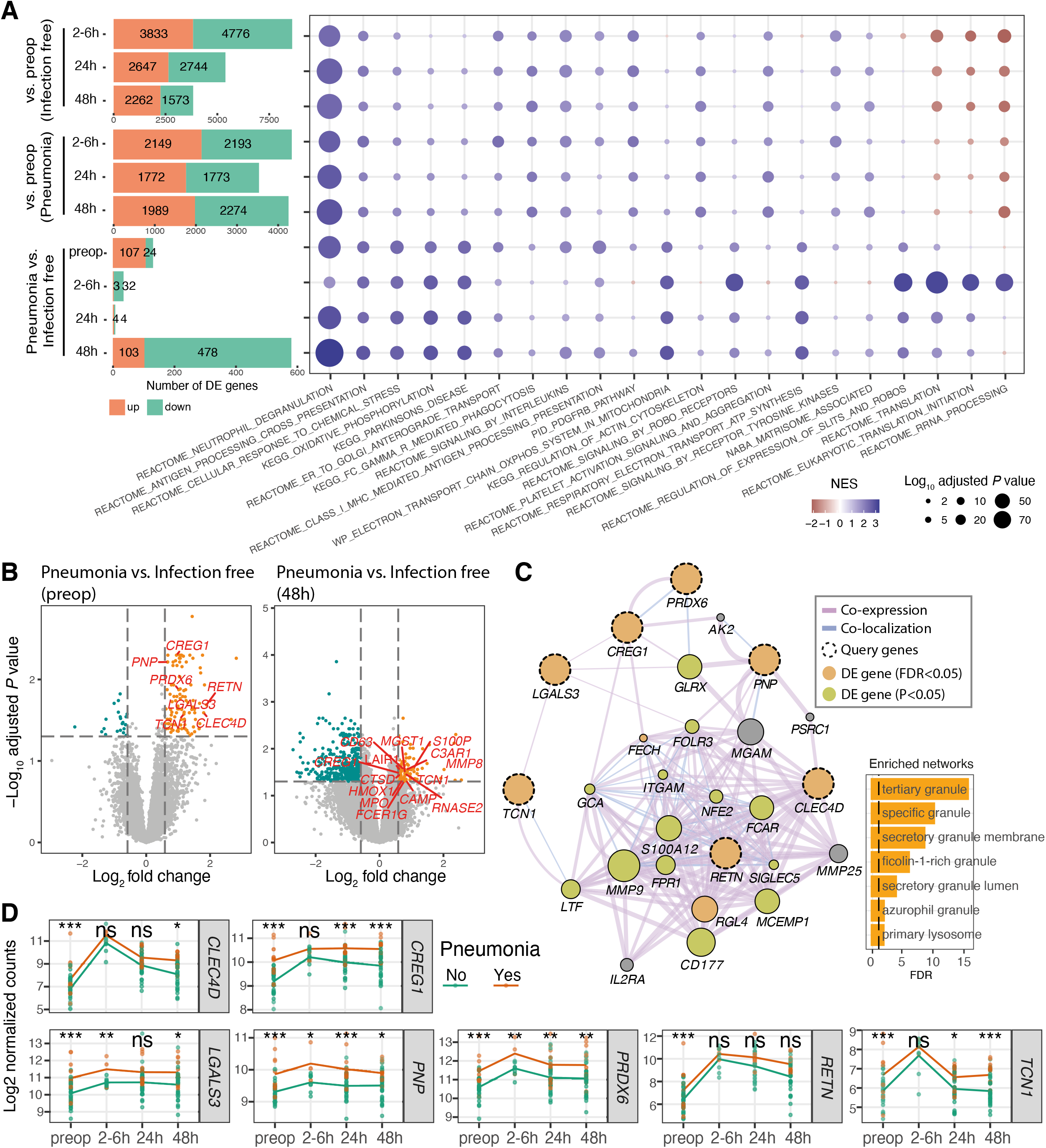
Neutrophilic inflammation may predispose patients to postoperative pneumonia. (A). Bar plots outlining the number of differentially expressed genes in each contrast as indicated on the y axis (left panel), and a heatmap (right panel) displaying the enrichment of MSigDB canonical pathways using fgsea (see **Supplementary Methods**). The top 5 pathways in each contrast were selected for visualisation. (B). Volcano plot showing differentially expressed genes in preoperative samples (left panel) and 48h samples (right panel). -Log_10_ adjusted *P* values are plotted against the log_2_ fold change of the gene expression (pneumonia vs. infection-free). The horizontal dashed lines represent the FDR-adjusted *P* value of 0.05. The differentially expressed genes are coloured in orange (up-regulated) and cyan (down-regulated). The differentially expressed genes involved in neutrophil degranulation and/or myeloid leucocyte mediated immunity pathway are labelled and highlighted in red. (C) Network showing the interactions between seven differentially expressed genes (dashed circles; highlighted in Fig. 3B left panel) with their related genes using GeneMANIA with a default setting. The purple edge colour represents co-expression, and blue edge represents co-localization. The edge width indicates the interaction weight. The size of the nodes for the related genes indicates the interaction score assessed by GeneMANIA. The bar chart showing the enriched GO terms assessed by GeneMANIA using the network genes (D). The dynamic changes of the seven differentially expressed genes comparing pneumonia patients (in orange) to infection-free patients (in cyan) across different time-points. * *P* < 0.05, ** *P* < 0.01, *** *P* < 0.001, *n.s*. = not significant.

Gene set enrichment analysis (see **Supplementary Methods**) was performed to explore whether functional gene sets coincident with the acute transcriptomic changes following major abdominal surgery were more evident in those who developed pneumonia compared to those remaining infection-free. These differentially expressed genes were enriched for immune and inflammatory pathways and GO terms (**Fig. 4A right panel & E4A**; **Supplementary Table E6**). Significant neutrophil (**Fig. 3A**) and myeloid mediated immunity (**Fig. E3A**) enrichment was observed in preoperative and postoperative samples obtained from those who subsequently developed pneumonia compared to those remaining infection free (Normalised Enrichment Score (NES)=2.7, adjusted *P*=5.1E-42 for neutrophil degranulation in **Fig. 4A**; NES=2.6, adjusted *P*=5.6E-39 for myeloid leucocyte mediated immunity in **Fig. E3A**). Importantly, this dynamic functional enrichment of gene sets persisted in all previously described contrasts when adjusted for cell proportions of neutrophils, monocytes and lymphocytes (**Fig. E3B-C**).

As immature neutrophils have recently been implicated as drivers of the poor outcome SRS1 state (20), we specifically investigated genes involved in neutrophil degranulation and innate immunity and found seven of these genes were differentially expressed in preoperative samples (**Fig. 4B** left panel; highlighted in red). These genes were all upregulated in both preoperative and 48h samples from patients later developing pneumonia when compared to samples from those remaining infection-free (**Fig. 4B**). As expected, we found that these seven genes form a co-expression/localisation network involved in neutrophil maturation and activation (**Fig. 4C**), with the increased expression persisting in postoperative samples from patients later developing pneumonia (**Fig. 4D**).

### A preoperative signature composed of genes involved in neutrophil degranulation is associated with the development of postoperative pneumonia

To assess the performance of the seven neutrophil-related gene signature we had identified, k-means clustering analysis was performed across the preoperative samples (**Fig. 5A**). Two k-means clusters were extracted and analysed to detect if cluster assignment could distinguish between those who did or did not develop pneumonia. All patients in cluster 2 (5 out of 5) developed pneumonia compared to 6 out of 34 patients in cluster 1 (*P* = 0.00092; **Fig. 5A-B**). Higher expression of these seven genes was also associated with postoperative pneumonia at later time points (24 and 48hrs) but not in the acute phase (2-6hr) (**Fig. 5B**). Similarly, when patient clusters were defined with hierarchical clustering, two distinct clusters were identified (**Fig. E4)**. Both pre-operatively and at 48hrs, higher proportions of patients in cluster 2 developed pneumonia (**Fig. 5C**).

**Figure 5.**
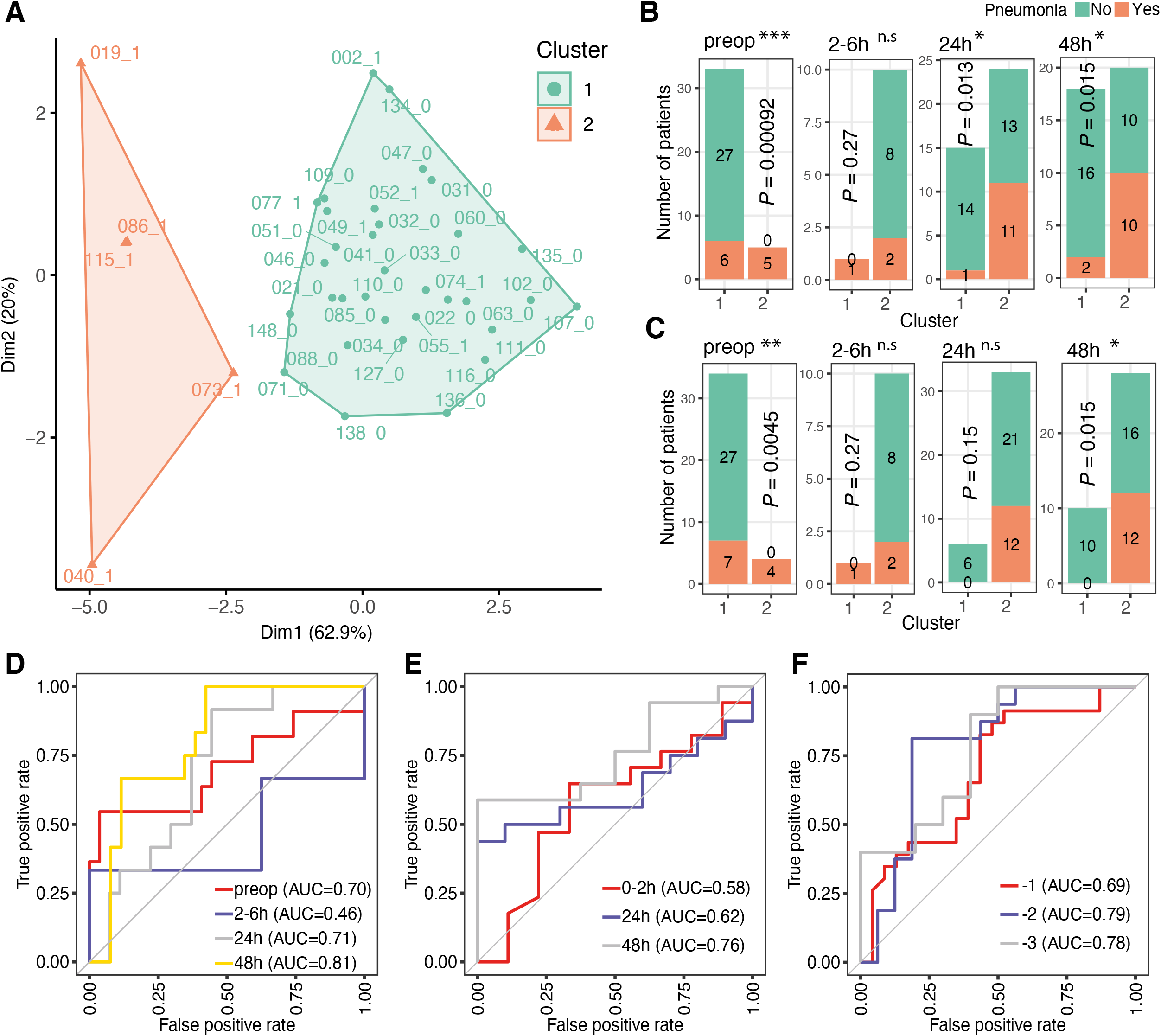
Predictive performance of a signature composed of genes involved in neutrophil degranulation for detection of postoperative infections. (A). Cluster plot showing K-means analysis of preoperative samples with the proportion of variance explained by each component shown. Two major clusters were highlighted in orange and cyan. The patient IDs together with the clinical status (1: pneumonia; 0: infection-free) were shown next to each dots. (B-C). Bar plots of the number of K-means (B) or hierarchical (C) cluster1 and cluster2 patients who did (in orange) or did not developed pneumonia (in cyan) across different time-points. *P* values were calculated by two-tailed Fisher’s exact test. * *P* < 0.05, ** *P* < 0.01, *** *P* < 0.001, *n.s*. = not significant. (D-F). ROC curves of prediction models constructed using the seven neutrophil-related gene expressions in the BIONIC cohort (D); a cohort with critically injured patients (E) (8) and a cohort with patients who develop postoperative infection up to three days prior to clinical diagnosis (F) (26).

Next the performance of the seven neutrophil-related gene signature in predicting patient outcomes was evaluated. The presence or absence of pneumonia was used as the outcome and models were constructed using the random forest method, based on the gene expression levels at each time-point (see **Supplementary Methods**). This gene signature achieved an AUC of 0.70, 0.71 and 0.81 preoperatively, at 24hrs and 48hrs respectively (**Fig. 5D**) but showed poor predictive performance 2-6hrs postoperatively.

Validation was performed by comparison of our data to two publicly available microarray datasets (8, 26) (see **Supplementary Methods**). In a cohort of critically injured polytrauma patients (8) whole blood transcriptome was analysed at 3 different time-points; the acute phase (0-2hr; within 2hrs of traumatic injury), 24 and 72hrs after injury. Good performance was observed using our seven neutrophil-related gene signature in predicting infection with AUC of 0.76 at 72hrs, but not at the acute phase time-point (0-2hr; AUC=0.58) (**Fig. 5E**). In an independent prospective cohort of elective surgery patients, which included patients who later developed a postoperative infection (26) our seven neutrophil-related gene signature also showed good predictive performance, up to three days prior to clinical diagnosis (26) (AUC 0.78 at day -3, 0.79 day -2 and 0.69 day -1; **Fig. 5F**).

### Predictive performance of a quantitative transcriptomic sepsis response score for the early identification of postoperative pneumonia

The results of our analysis indicate potential predictive utility of preoperative whole blood transcriptomics involving neutrophil function. Given our recent work developing a quantitative transcriptomic sepsis response score (18) (SRSq; ranging from 0 to 1) that in part reflects neutrophil dysfunction and altered granulopoiesis (20) with a greater risk of adverse outcomes (18), we hypothesised that SRSq might be associated with postoperative pneumonia risk. We found that SRSq scores were increased in postoperative samples relative to preoperative samples (**Fig. 6A**) and that SRSq was higher at 48hrs in those patients who ultimately developed pneumonia compared to those remaining infection-free (*P*=0.045, **Fig. 6B**). Furthermore, SRSq predictive performance at 48hrs postoperatively showed an AUC of 0.69 (**Fig. 6C)**. It is important to note that SRSq differences were not evident preoperatively or at earlier postoperative time-points (**Fig. 6B and 6C**). Using an extended (19 gene) methodology to calculate SRSq (18), we observed more significant differences between pneumonia and infection-free patients at 48hr (*P*=0.0095, **Fig. E5**).

**Figure 6.**
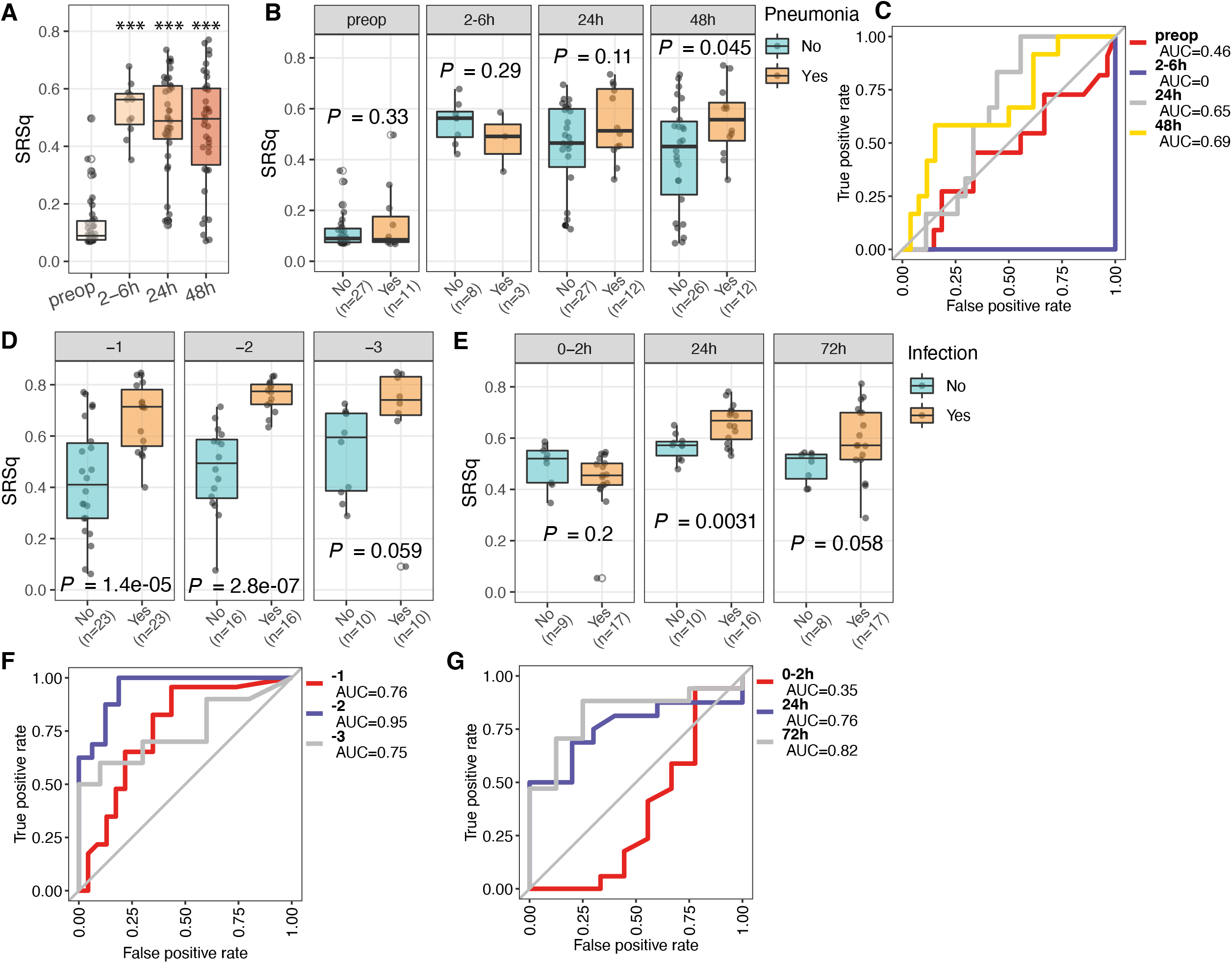
Predictive performance of the transcriptomic sepsis response score for detection of postoperative infection. (A-B). Box plots of the SRSq scores for samples in each time-point (A) or for samples from patients with (orange) or without (cyan) pneumonia (B). SRSq was quantified using a pre-defined and previously validated seven gene set through SepstratifieR package (18). *P* value was determined using a linear model, *** *P* < 0.001. (C). Receiver Operating Characteristic (ROC) curves of prediction models constructed using SRSq scores for samples in the BIONIC cohort. AUC: Area under the (ROC) Curve. (D). Bar plots showing the SRSq scores in a cohort with patients who develop postoperative infection up to three days prior to clinical diagnosis assayed at days -1 (n=46), -2 (n=32) and -3 (n=20) (26). (E). Bar plots showing the SRSq scores in a cohort with critically injured patients (8) assayed at 0-2h (n=26), 24h (n=26), and 72h (n=25) post injury. (F-G). ROC curves showing the performance of SRSq scores in predicting infection in the two cohorts as described above.

To validate these findings, we interrogated published data from the independent cohort of elective surgery patients (26) and found that SRSq could distinguish those who later developed infection from matched patients remaining infection-free at one (*P*=1.4×10^−5^) and at two (*P*=2.8×10^−7^) days prior to the clinical diagnosis of infection (**Fig. 6D**). In critically injured polytrauma patients (8), we found that SRSq scores differed significantly between patients who subsequently developed infection compared to those remaining infection-free, but only at later time-points (after 24hr) (**Fig. 6E)**. Consistently, SRSq performed well as a predictor of infection in both cohorts (**Fig. 6F-G**; AUC of 0.76, 0.95 and 0.75 at day -1, -2 and -3 respectively in the prospective cohort of elective surgery patients; AUC of 0.76 and 0.82 at 24hrs and 48hrs respectively in the polytrauma patient cohort), but not at the acute phase time-point (**Fig. 6G;** 0-2hr; AUC=0.35).

## Discussion

In this prospective observational transcriptomic analysis of patients undergoing major abdominal surgery, pathway analysis demonstrated postoperative innate-immune activation, including changes in neutrophil degranulation and TLR signalling alongside adaptive immune suppression, characterised by the downregulation of T-cell receptor signalling and expression of T-cell exhaustion markers. Bulk-RNA cell type deconvolution revealed increases in circulating HSPCs and S100A8/9-high neutrophils alongside reductions in naïve CD4 T cells in patients later diagnosed with postoperative pneumonia, while gene-set enrichment analysis revealed up-regulation of neutrophil degranulation and myeloid-leucocyte mediated immunity. Importantly, these cell composition changes and pathway enrichments are evident preoperatively in those later suffering pneumonia. SRSq scores, reflecting relative immunosuppression and a more deranged host response, increased postoperatively relative to paired preoperative samples and at 48hrs scores differed significantly between patients who subsequently developed a nosocomial pneumonia and those remaining infection-free. The expression of seven genes, characteristic of neutrophil maturation and activation showed good predictive performance preoperatively and 48hr postoperatively in identifying patients later developing pneumonia. Analysis of this novel gene-set and the SRSq score in independent cohorts undergoing major surgery or following polytrauma showed good predictive performance in identifying patients who later developed nosocomial infection.

A recent analysis of perioperative microarray data demonstrated a series of differing gene signatures, derived by statistical means rather than biological plausibility, that distinguish between patients who develop postoperative infection up to three days prior to clinical diagnosis (26). This study (26), previous studies describing the utility of SRS/SRSq in infectious diseases (16-18), and the gene signatures described here involving neutrophil and myeloid mediated immunity, emphasise the informativeness of transcriptomic patterns for delineating biological function and pathway activation in acute illness. They also highlight the opportunity to develop point-of-care testing for relatively small combinations of genes, over time for prognostication and targeted intervention (26).

Examining the content of the gene sets and SRS/SRSq implicates mature, S100A8/9 high, degranulating, IL1R2^+^, immature and PADI4^+^ neutrophils as the underlying drivers of SRS membership/SRSq (20). Our signature is based on differentially expressed genes characteristic of neutrophil maturation and activation, rather than an unsupervised analysis. S100A8/9-high neutrophils, identified from bulk-RNA deconvolution, are known to act as an endogenous ligand for TLR-4 (27) and the receptor for advanced glycation end product (RAGE) (28), and have been negatively correlated with sepsis recovery (20), as well as being implicated in SARS □ CoV □ 2 induced viral pneumonitis (29). We observed a heightened abundance of these two cell types in both the preoperative and postoperative samples of pneumonia patients, implying that the functional effects of S100A8/9-high neutrophils may predispose to postoperative pneumonia.

Preoperative neutrophil degranulation is evident in those with an increased risk of nosocomial pneumonia. The conventional view of neutrophils as homogeneous, short-lived cells playing a passive role in coordinating inflammation is being revisited (30). During malignancy deranged myelopoiesis drives the expansion of polymorphonuclear (PMN) myeloid-derived suppressor cells from the bone marrow, impairing immune surveillance (31). Elevated concentrations of these heterogeneous, poorly defined neutrophil-like populations are associated with a worse prognosis (32, 33). In this context, neutrophil degranulation has been implicated in tumour progression and the facilitation of metastatic dissemination (34) and may identify patients with a more severe disease process than is apparent when assessed by conventional preoperative clinical scoring. Indeed, elevations in NLR are associated with worse survival in many solid tumours (35).

Following lessons learned from cohort heterogenicity in the sepsis literature (36) pneumonia (23) was selected rather than *all cause* nosocomial infection as an enrichment strategy (37) ensuring our clinical endpoint was as robust and homogeneous as possible. We acknowledge that inter-individual variation in the perioperative immune response will play a role in susceptibility to other nosocomial infectious complications such as intra-abdominal abscess, secondary to anastomotic breakdown or surgical site infection (38), but this signal will be diluted by the contribution of surgical complications to these types of infection.

The overall effects of both volatile and intravenous anaesthesia are thought to be immune suppressive (6, 7). However, patients recruited to this study received a consistent anaesthetic strategy, with the most common combination being neuraxial analgesia (either an epidural (59%) or a single-shot intrathecal injection (8%)) with an intravenous anaesthetic induction and maintenance with a volatile anaesthetic (sevoflurane or desflurane). More recently, there has been an interest in the influence of anaesthetic choice on cancer outcomes (39). Cancer associated immunological changes are characterised by T-cell exhaustion, the presence of circulating myeloid derived suppressor and T_reg_ cells and are thereby predominantly immunosuppressive (14, 15). These effects of cancer on immune function may have influenced the differential transcriptomic patterns described in this study.

This study does have limitations. Whole-blood bulk RNA sequencing provides an accessible, unbiased snapshot of the circulating transcriptome. As blood is a dynamic compartment, its circulating cellular make-up is expected to represent rapid alterations in immune function. However, sampling only blood may miss compartmentalised responses that can occur in sterile insults (40). Peripheral myeloid expansion occurs with malignancy (41) and following physiological stress (30) meaning that comparisons between quiescent preoperative samples and postoperative samples should be interpreted with caution, despite adjusting for paired differential leukocyte cell-counts.

This study demonstrates the potential of transcriptomics to identify preoperatively patients who are at increased risk of developing a postoperative pneumonia. Sepsis response signature scores postoperatively and preoperative transcriptomic signatures of neutrophil degranulation may be useful predictors of the subsequent risk of pneumonia. These findings raise the potential for the preoperative identification of patients susceptible to developing pneumonia, bringing us closer to a precision medicine approach to the prevention of postoperative infections. However, as this study was intended to be hypothesis generating, findings will require replication. Additional work specifically focusing on neutrophil heterogeneity using microscopy and single-cell analysis techniques is warranted. While the identification of the shared pathways between the immune response to malignancy and susceptibility to infection, will be beneficial in this high-risk perioperative cohort.

## Supporting information

Supplementary_Table_S1_BIONIC_cohort_meta

Supplementary_Table_S2_DE.genes.2-6h.vs.preop

Supplementary_Table_S3_pathway.enrichment

Supplementary_Table_S4_difference.in.cell.abundance

Supplementary_Table_S5_DE.genes.Pneumonia_Yes_vs_No

Supplementary_Table_S6_GSEA.results

## Data Availability

All data produced in the present study are available upon reasonable request to the authors

## Acknowledgements

The authors are grateful to the Adult Critical Care Research Team of the Royal London Hospital, Barts Health NHS Trust for data collection and blood sampling.

## Author contributions

Professors Gordon and Knight had full access to all the data in the study, take responsibility for the integrity of the data and the accuracy of the data analysis and had final responsibility for the final decision to submit for publication.

*Study concept and design:* HDT, PZ, MJO’D, CJH, JCK … ACG.

*Acquisition, analysis, or interpretation of data:* All authors.

*Drafting of the manuscript*: HDT, PZ, CJH, JCK … ACG.

*Critical revision of the manuscript for important intellectual content:* All authors.

*Statistical Analysis:* HDT, PZ.

*Obtained funding:* HDT, PZ, CJH, JCK … ACG.

*Administrative, technical, or material support:* HDT, PZ, ERL, YM, JPW, AA, AJK, CGG … EC-G.

All authors read the final draft of the manuscript and confirm to the accuracy and integrity of the work.

## Funding

National Institute for Health Research (NIHR) Academic Clinical Fellowship (ACF-2019-21-011) (HDT), Clinical Lectureship (CL-2022-21-005) (HDT) and Research Professorship (RP-2015-06-018) (ACG), The National Institute of Academic Anaesthesia (NIAA) BJA/RCoA – Project Grant (WKR0-2020-0019) (HDT, ACG and JCK), Medical Research Council (MR/V002503/1) (JCK and EED), Wellcome Trust Investigator Award (204969/Z/16/Z) (JCK), Wellcome Trust core funding to the Wellcome Sanger Institute (Grant numbers 206194 and 220540/Z/20/A), Wellcome Trust Grants (090532/Z/09/Z and 203141/Z/16/Z) to core facilities Wellcome Centre for Human Genetics, The Chinese Academy of Medical Sciences (CAMS) Innovation Fund for Medical Science (CIFMS) (grant number: 2018-I2M-2-002) (JCK, PZ), NIHR Oxford Biomedical Research Centre (JCK) and NIHR Imperial Biomedical Research Centre (HDT, ACG).

## Declarations

Professor Rupert M Pearse declares research grants and/or honoraria from Edwards Lifesciences, Intersurgical and GlaxoSmithkline and Professor Anthony C Gordon FMedSci declares consulting fees from AstraZeneca, both unrelated to this project. All other authors report no conflict of interest.

## BIONIC IDs

These were not known to anyone outside the research group and cannot reveal the identity of the study subjects.

## Supplementary Figure Legends

**Figure E1.**
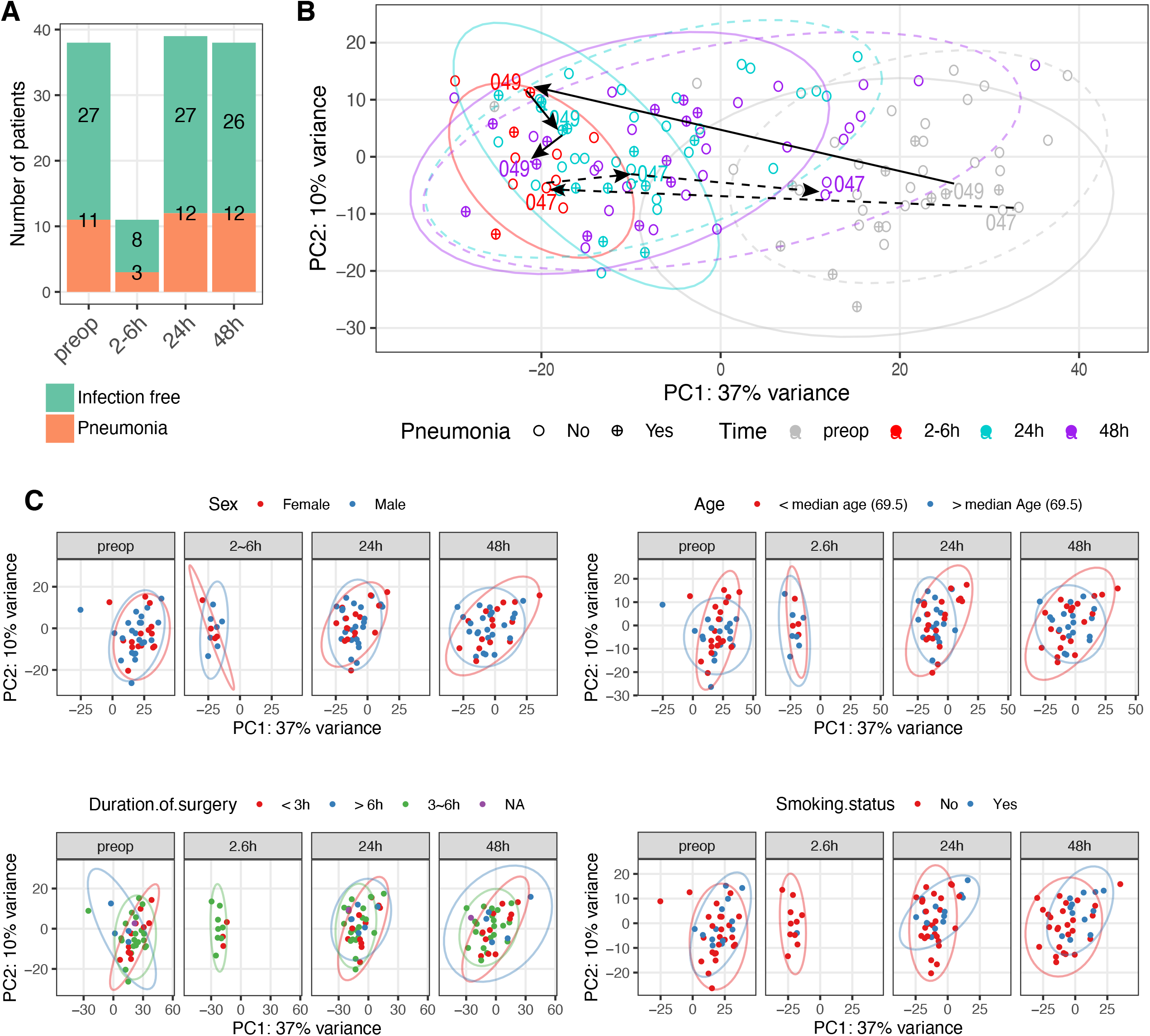
(A). Bar plot outlining the number of patients sampled for RNA sequencing at each timepoint. (B). Principal component analysis of gene expression in samples preoperatively and postoperatively in 2-6, 24 and 48hrs. Each dot represents a sample, labelled with patient ID. Colours indicate different time-points. Shapes indicate the clinical status of patients postoperatively (circle: infection-free; circle plus: pneumonia). Solid arrow represents the trajectory of patient (ID: 049) later suffering pneumonia compared to the dashed line depicting a patient (ID: 047) remaining infection free. (C). Principal component analysis of gene expression in samples preoperatively and postoperatively in 2-6, 24 and 48hrs. Each dot represents a sample and is coloured with either sex (upper left panel), age (upper right), duration of surgery (lower left) or smoking status (lower right).

**Figure E2.**
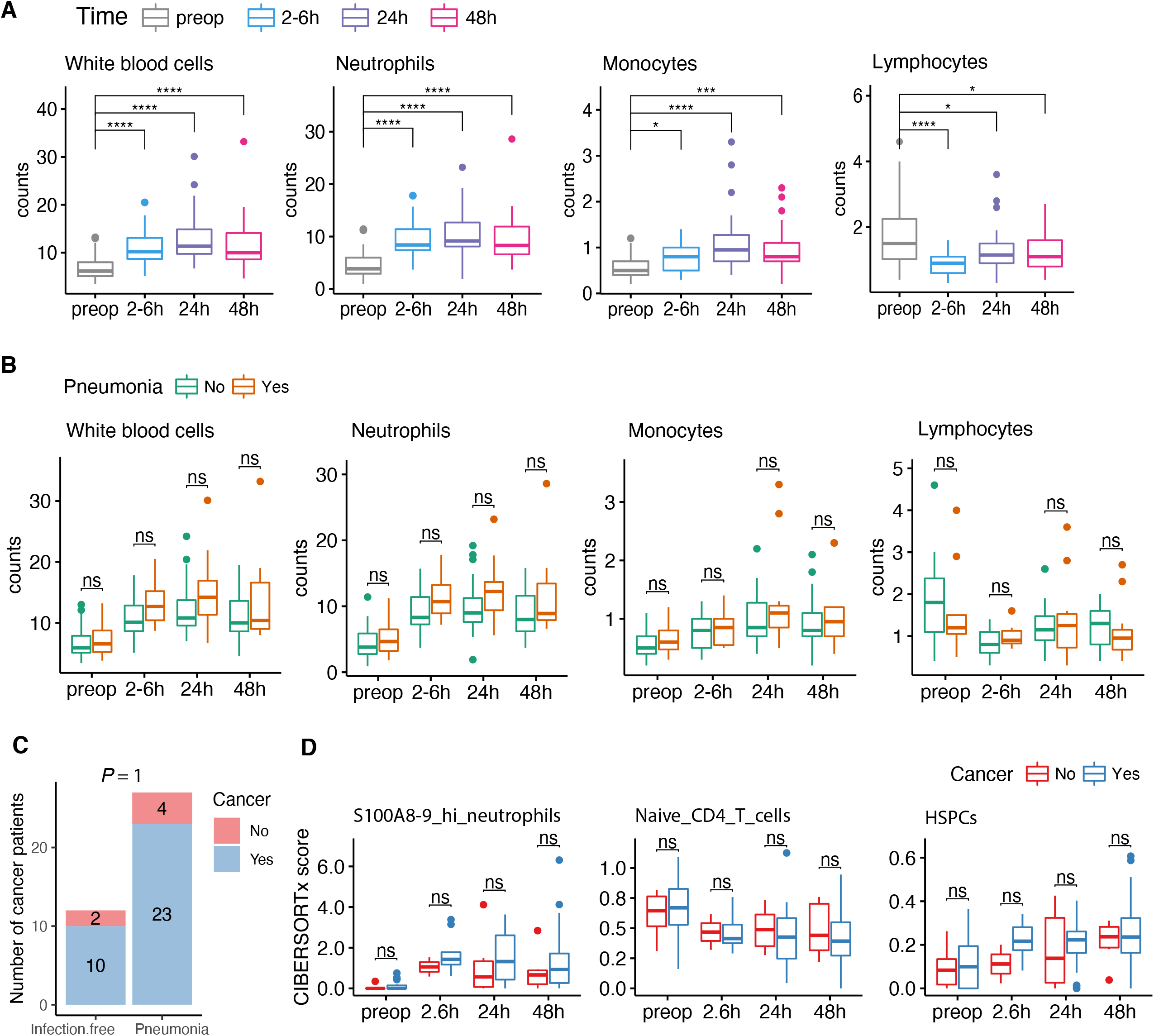
(A). Cell counts (×10^9^/L) in samples across different time-points. *P* value was calculated by Wilcoxon signed-rank test. * *P* < 0.05, *** *P* < 0.001, **** *P* < 0.0001, *n.s*. = not significant. (B). Cell counts (×10^9^/L) in samples from patients who did (orange) or did not develop pneumonia (cyan) across different time-points. *P* value was calculated by Wilcoxon signed-rank test. *n.s*. = not significant. (C). Bar plot outlining the number of cancer patients who did or did not develop pneumonia. *P* values were calculated by two-tailed Fisher’s exact test. (D). Box plots of CIBERSORTx absolute scores of indicated cell types in samples from patients with (red) or without cancers (blue) across different time-points. *P* value was calculated by Wilcoxon signed-rank test. *n.s*. = not significant.

**Figure E3.**
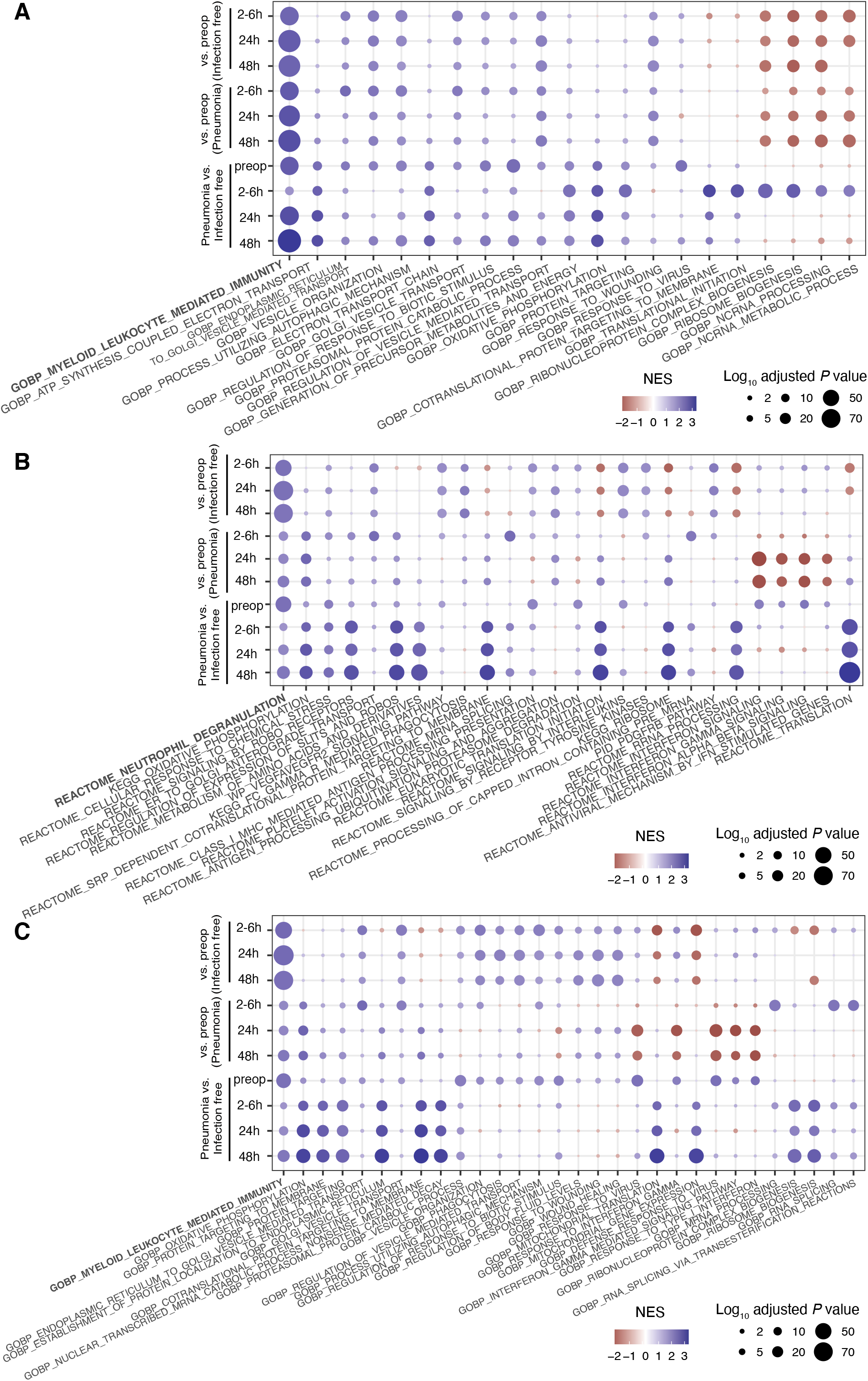
(A-C). Heatmaps illustrate the enriched pathways/terms using gene set enrichment analysis. The top 5 enriched biological process GO terms (A), Canonical pathways corrected for cell proportions of neutrophils, monocytes and lymphocytes (B) or biological process GO terms corrected for cell proportions of neutrophils, monocytes and lymphocytes (C), in each contrast as indicated on the y axis were merged for visualisation.

**Figure E4.**
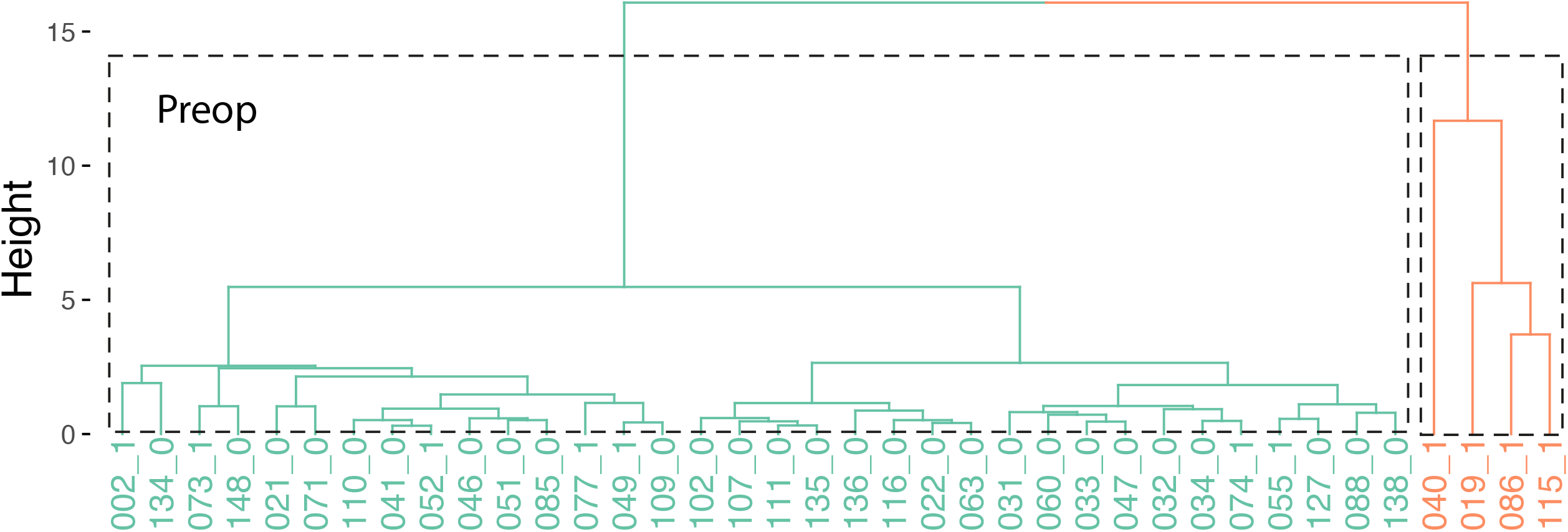
Dendrogram showing agglomerative hierarchical clusters in preoperative samples. The height on the y-axis represents the distance between two clusters. Two major clusters are highlighted in orange and cyan. The patient IDs together with the clinical status (1: pneumonia; 0: infection-free) are shown at the bottom of the dendrogram.

**Figure E5.**
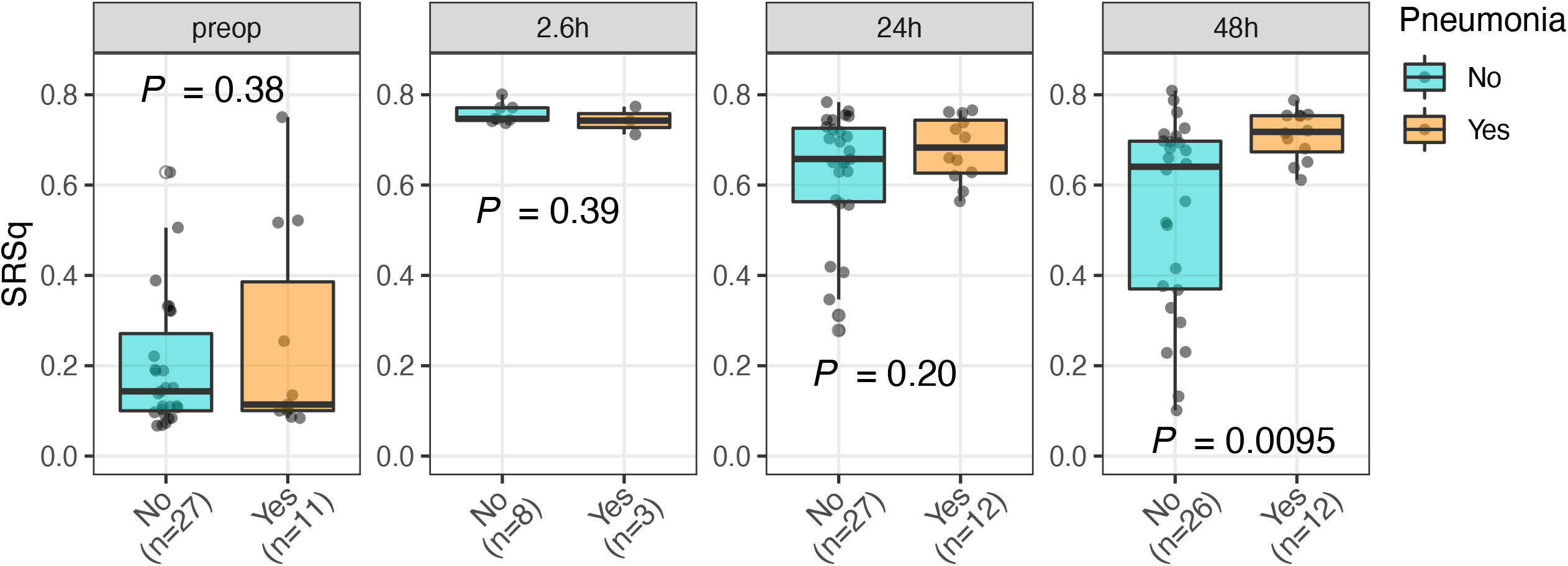
Dot plots of the SRSq scores for samples from patients with (orange) or without (cyan) pneumonia. SRSq was quantified using the extended 19 gene signature to show robustness by predictive gene set (18). *P* value was determined using a linear model.

## Notes

### Funding Statement

This study was supported in part from the National Institute for Health Research, the National Institute of Academic Anaesthesia, the Medical Research Council, the Wellcome Trust, and the Chinese Academy of Medical Sciences Innovation Fund for Medical Science.

### Author Declarations

East Midlands Nottingham 2 Research Ethics Committee gave ethical approval for this work.

