## Supplementary Methods for "A transcriptomic approach to understand patient susceptibility to pneumonia after abdominal surgery"

**Study details and ethics**

The BIONIC (Biomarker based Identification Of Nosocomial Infective Complications) study was a prospective observational study. One hundred and fifty consecutive patients undergoing elective major abdominal surgery patients were recruited between April 2015 and August 2016 at the Royal London Hospital. Ethics approval was granted by the East Midlands – Nottingham 2 Research Ethics Committee (14/EM/1223).

**Inclusion and exclusion criteria**

The inclusion criteria was age 45 or over, undergoing scheduled surgery involving the gastrointestinal tract, requiring a general anaesthetic and at least an overnight hospital stay

The exclusion criteria were refused consent and emergency surgery. Every patient on a weekday elective operating list was screened. Eligible patients were then approached for written, informed consent. Entry into the study did not influence clinical management. All patients received standardised perioperative prophylactic antibiotic therapy.

**Data Collection**

Data were collected, in real time, on each patient until hospital discharge and included information on co-morbidities, American Society of Anesthesiology (ASA) physical status classification, indication for surgery, cancer staging and diagnosis, duration of the procedure, planned postoperative intensive care unit admission and in-hospital mortality. Patients were examined daily for the presence of infection. Definitions of infection were agreed prospectively by the investigators and were based on the Centre for Disease Control and Prevention (CDC) criteria (1).

**Blood sampling**

Blood samples were drawn immediately before induction of anaesthesia (preoperatively) and then between 2-6, and at 24 and 48hrs following the operation. Lack of availability of research staff impinged on the practicalities of obtaining the 2-6hr sample so sampling at this time was discontinued after the first 50 patients. PAXGene (Qiagen, USA) RNA tubes were collected and stored at -80^o^C until analysis. A paired EDTA sample was drawn alongside each PAXGene tube and a differential leukocyte count was performed by hospital laboratory staff, using a Sysmex SE2100 Analyser (Sysmex, Milton Keynes, UK).

**RNA extraction**

RNA was collected in PAXGene (Qiagen, USA) tubes and extracted using the PAXGene RNA extraction kit, with a DNase step to remove contaminating DNA. The integrity and quality of the total RNA were assessed on an Agilent 2100 Bioanalyzer (Agilent Technologies, Germany) by means of the RNA 6000 Nano Assay (Agilent Technologies, Germany). Globin and ribosomal RNA were removed via a Ribo-zero kit (Illumina, USA) and library preparation was carried out using the TruSeq Stranded Total RNA Library Prep Kit (Illumina, USA). Next generation sequencing was performed on the NovaSeq sequencing platform (Illumina, USA) at the Wellcome Centre for Human Genetics (WHG) in Oxford.

**RNA-Sequencing data analysis**

RNA sequencing read quality was assessed by FastQC (v0.11.9), with reads trimmed using Trim Galore (v0.6.2) and mapped to human genome primary assembly GRCh38 using STAR (v2.7.3a) (2) in multi-sample 2-pass mode and based on the ENCODE recommended parameters. The aligned BAM files were quality checked via RNA-SeQC (v2.3.6), and then used to determine the gene counts through GENCODE annotations (release 34) and featureCounts (v1.6.4). After RNA-seq mapping quality assessment, one sample (ID:133 preop) with a low mapping rate and high proportions of duplicates was flagged as poor quality and removed from further analysis. Potential sample swaps were checked using CrosscheckFingerprints function from Picard (v 2.21.1) (3). For gene differential expression analysis, the raw read counts were used as input into the R package DESeq2(4) (v1.28.1) for pair-wise comparisons. Genes that had <10 reads mapped in >90% of the samples and the sex chromosome genes were filtered, retaining 18,653 genes for downstream analysis. Genes with fold change >1.5 and FDR <0.05, as per condition, were considered to be differentially expressed.

The raw sequencing datasets and processed results generated for this study are available upon request and will be made publicly available upon publication.

All analyses were performed using the R software (v4.0.3).

**Validation datasets**

Trauma cohort: (5) Files with the normalised gene expression data (microarray) and the clinical data were retrieved from <https://github.com/C4TS/HyperacutePhase>

Perioperative cohort: (6) Files with the normalised gene expression data (microarray) and the clinical data (batch1) were retrieved from <https://data.mendeley.com/datasets/rhc5s6zj88/3>

**Cell type deconvolution of bulk RNA-seq data**

Cell-type deconvolution was performed with CIBERSORTx (7) using a reference panel derived from a single cell RNA-seq cohort of 26 sepsis patients including 9 convalescent samples, 6 healthy donors and 7 post-cardiac bypass surgery patients (8). A signature matrix was built by the Create Signature Matrix analysis module with parameters min. expression = 0.25, replicates = 100 and sampling = 0.5. The CIBERSORTx (7) absolute scores of each cell type in bulk samples were then obtained using the mixture file (Bulk RNAseq count matrix normalised by DESeq2 (4)), the signature matrix derived from single cell RNA-seq, the single cell reference matrix for S-mode batch correction and with 100 permutations via the Impute Cell Fractions analysis module.

**Gene enrichment and network analysis**

The hypergeometric pathway enrichment analysis was performed as described previously (9). Differentially expressed genes, background genes expressed in this dataset (n=18,653), and the Reactome gene sets downloaded from Molecular Signatures Database (MSigDB; v2022.1) were used for analysis. Significance was determined using PHYPER function as implemented in R and multiple hypotheses testing by Benjamini–Hochberg correction. Gene-set enrichment analysis (GSEA) was performed using R package fgsea and MSigDB v7.4.1 including Canonical pathways (KEGG, BIOCARTA, REACTOME, PID and WikiPathways) and Gene Ontology (GO) terms. GSEA was carried out separately for each contrast, with genes ranked by both p-value and fold change/direction (-log_10_[*P-*value] x sign[log_2_FoldChange]). The top 5 enriched pathways in each contrast were selected for visualisation. GeneMania (v3.5.2) (10) and Cytoscape (v3.9.0) (11) were used to query and visualise the gene networks.

**Clustering analysis**

Patient clusters were defined by hierarchical agglomerative clustering based on a similarity measure (Euclidean distance) and Ward's method or k-means (Hartigan-Wong algorithm). The R package factoextra (v1.0.7) was used for cluster visualisation.

**Machine learning**

We evaluated the performance of our gene signature with three repeats of 10-fold cross-validation and a random forest approach as implemented in the R package caret (v6.0.92). AUC and ROC curves of the models were calculated and plotted using the R package MLeval (v0.3).
